# CausalDRIFT: Causal Dimensionality Reduction via Inference of Feature Treatments for Robust Healthcare Machine Learning

**DOI:** 10.1101/2025.07.10.25331298

**Authors:** Kazi Sakib Hasan, Jarin Akter Mou

## Abstract

High-dimensional medical datasets present challenges in feature selection, where traditional methods often prioritize spurious correlations over causally relevant variables, compromising model interpretability and clinical utility. We introduce CausalDRIFT, a causal feature selection algorithm grounded in the Frisch-Waugh-Lovell theorem and Double Machine Learning, which estimates the Average Treatment Effect (ATE) of each feature on clinical outcomes while adjusting for confounders. We evaluated CausalDRIFT against seven baseline methods (PCA, ICA, Elastic Net, RFE, etc.) across four datasets (Heart Disease, Diabetes, Breast Cancer, and PCOS) using XGBoost classifiers, with performance metrics including accuracy, precision, recall, and F1-score. CausalDRIFT achieved competitive performance, notably excelling in datasets with strong causal structure (e.g., 90% accuracy and F1-score of 0.90 on PCOS, outperforming most other methods). It demonstrated superior consistency (lowest standard deviation: 1.19 in Breast Cancer, lowest recall spread in Heart Disease) and robustness to confounding, though it traded marginal predictive gains for interpretability in correlation-dominated datasets. CausalDRIFT excels in high-dimensional, low-sample size (HDLSS) settings, such as the Breast Cancer dataset (569×32), where it achieves 93.9% accuracy and 91.8% F1-score. Statistical analysis (ANOVA, Tukey HSD) confirmed its recall performance was non-inferior to top methods (all p > 0.15), while unsupervised techniques like ICA significantly underperformed (p < 0.05). CausalDRIFT bridges the gap between causal inference and scalable feature selection, offering clinically interpretable and generalizable models. Its ability to prioritize causally actionable features is critical for high-stakes decision-making, and it makes it a promising tool for healthcare AI, particularly in settings like PCOS where etiology is complex.

## 1 Introduction

In the era of data-driven medicine, healthcare systems are increasingly leveraging large-scale datasets encompassing electronic health records (EHRs), clinical notes, laboratory results, genomic profiles, and medical imaging. These datasets are often high-dimensional, containing hundreds to thousands of variables per patient. While this abundance of information offers unprecedented opportunities for predictive modeling and personalized care, it also presents substantial challenges. Chief among them is the curse of dimensionality, wherein the performance of machine learning models deteriorates due to the presence of noisy, redundant, or irrelevant features [1]. This not only increases computational burden but also amplifies the risk of overfitting, making models less generalizable across patient populations. Traditional feature selection and dimensionality reduction techniques have been developed to address these issues. Techniques such as Principal Component Analysis (PCA), Independent Component Analysis (ICA), and autoencoders are commonly employed to reduce the feature space by transforming or selecting variables based on statistical properties like variance or independence [2] [3]. The RFE (Recursive Feature Elimination) is another statistical method that leverages the generalization ability of Support Vector Machines (SVMs) to identify and retain the most important features while eliminating less relevant ones [4]. There is also correlation-based feature selection, which assumes that highly correlated features with the class are from relatively better feature sets than those of uncorrelated [5]. However, in healthcare applications, where decisions can significantly impact patient outcomes, relying solely on statistical associations is often insufficient. Correlation-based reductions may preserve variables that are only spuriously associated with the outcome, potentially leading to misleading interpretations and suboptimal clinical decisions [6]. Moreover, many high-dimensional healthcare datasets suffer from heterogeneity and hidden confounding, further complicating feature selection. For instance, lab test values might appear predictive of patient mortality but may actually reflect clinician decisions or institutional practices rather than intrinsic disease severity. In such settings, understanding whether a feature is causally relevant rather than merely predictive is of paramount importance [7]. Without causal reasoning, feature selection pipelines risk incorporating variables that are proxies or consequences of the outcome rather than true drivers, undermining both model robustness and interpretability. Thus, there is a pressing need for feature selection methods that go beyond correlational analysis and instead focus on uncovering the underlying causal structure of the data. Such methods can provide more robust, explainable, and transferable models—characteristics that are particularly critical in high-stakes domains like clinical decision support and treatment recommendation systems.

Feature selection and dimensionality reduction are foundational to building efficient and interpretable machine learning models, especially when dealing with high-dimensional datasets. In healthcare, popular methods include filter-based approaches (e.g., correlation coefficients), wrapper methods like Recursive Feature Elimination (RFE), and embedded techniques such as Elastic Net, which simultaneously perform regularization and feature selection [9]. Unsupervised transformations such as Principal Component Analysis (PCA) and Independent Component Analysis (ICA) are also widely employed to project data into lower-dimensional spaces while preserving statistical structure. Such techniques critically affect interpretability because they transform the original features into new components. It results in original and realistic feature loss, mixed and uncorrelated variables, and no feature-level explanation like regression coefficients [8]. However, despite their success in various domains, these techniques suffer from fundamental limitations when applied to healthcare data. Most notably, they are agnostic to causality, selecting or transforming features based on statistical association with the outcome rather than causal relevance. This distinction is not trivial, especially in clinical contexts where decisions informed by a model may directly affect patient care. For instance, a variable might exhibit a strong correlation with a clinical outcome but be downstream of the outcome or influenced by unmeasured confounders. In such cases, selecting this feature may lead to reverse causation or spurious inferences. For example, the administration of vasopressors may correlate with patient mortality in an ICU dataset, but it is not the cause of mortality, rather it is a response to patient deterioration. Traditional methods would incorrectly prioritize such variables, potentially compromising the interpretability and clinical trust in the model [10]. Furthermore, many of these techniques fail to account for the complex interactions and confounding structures that are prevalent in observational medical datasets. PCA and ICA, for example, construct linear combinations of original features without regard for the domain-specific meaning or intervention relevance of those features. This makes the resulting components difficult to interpret and inappropriate for applications where clinical explainability is essential. Elastic Net and other embedded methods can help reduce feature space while handling multicollinearity, but they still operate under purely associational frameworks. As such, they are sensitive to spurious correlations that may arise from dataset-specific quirks or unobserved biases, leading to models that are not generalizable or robust across clinical settings [11]. In summary, while traditional feature selection and dimensionality reduction methods offer valuable statistical tools, they lack the ability to distinguish between correlation and causation, which is a critical distinction in medical decision-making. There is a growing recognition in the machine learning and high-risk domains like healthcare communities that causal reasoning must be integrated into the modeling pipeline to ensure trustworthiness, fairness, and interpretability of clinical AI systems [12].

In clinical decision-making, the distinction between correlation and causation is not merely academic, rather it is foundational to safe and effective healthcare. While predictive models based on statistical associations can identify patterns in data, they often fail to explain why a relationship exists. In medicine, this explanatory gap can lead to interventions that are ineffective at best and harmful at worst. Therefore, the integration of causal reasoning into medical modeling is not only desirable but essential. Causal reasoning allows practitioners and algorithms alike to answer counterfactual questions: what is the average effect of treatment X on disease Y? For example, a high lactate level may be statistically associated with sepsis-related mortality, but treating lactate directly is not a clinically sound strategy, what matters is identifying and acting upon the cause of elevated lactate [13]. Predictive models trained on observational data without accounting for confounding and causal directionality can encode spurious associations. Such models may exploit shortcuts or proxy variables that are predictive in a given dataset but lack stability under distributional shifts [10]. This is especially dangerous in medicine, where patient demographics, institutional protocols, and measurement practices can vary widely between settings. By incorporating causal principles, models can become more robust, generalizable, and trustworthy. Furthermore, causal feature selection enhances interpretability, a key requirement for clinician adoption and regulatory approval. When a model selects features based on their causal relevance to an outcome, clinicians can more confidently understand and scrutinize its recommendations on the basis of Average Treatment Effect (ATE). Unlike latent factors generated by autoencoders or PCA components, causally selected features align more naturally with domain knowledge and biomedical mechanisms [14]. Importantly, the need for causal reasoning is amplified by the growing interest in precision medicine, where treatments are tailored to individual patients based on biomarkers and historical data. In this context, identifying which variables truly influence outcomes is not just useful, it is vital for ensuring that algorithms support, rather than hinder, personalized care strategies [15]. Therefore, predictive models in medicine must move beyond statistical association and embrace causal inference to achieve robustness, interpretability, and clinical relevance. Embedding causal reasoning into the feature selection stage, as proposed in this work, is a necessary step toward trustworthy and actionable machine learning in healthcare.

To overcome the limitations of purely associational feature selection methods, we introduce CausalDRIFT, a causal dimensionality reduction algorithm designed to identify features that exert a genuine causal influence on clinical outcomes. Unlike traditional techniques that rely on statistical correlation, CausalDRIFT draws on principles from causal inference to estimate how changes in individual features would affect outcomes, even in the presence of confounding. The key innovation behind CausalDRIFT lies in combining machine learning models for confounder adjustment with a simple causal effect estimation framework. For each feature, the method isolates the part that is not explained by other variables (its residual), does the same for the outcome, and then estimates the effect of the feature on the outcome via a clean regression between the two residuals. This approach is grounded in the Frisch-Waugh-Lovell (FWL) theorem, extended to non-linear settings using Double Machine Learning [16]. By applying this logic across all features, CausalDRIFT produces a ranked list of variables based on their estimated Average Treatment Effects (ATEs), which is a direct measure of their causal influence. Features with stronger causal effects are retained, while spurious or confounded ones are filtered out. This enables models to focus on variables that are more likely to reflect underlying mechanisms, rather than dataset-specific artifacts. In the context of medical data, this is especially valuable. Clinical datasets often contain noisy, proxy, or post-treatment variables that can mislead models. CausalDRIFT helps filter out such spurious correlations and surface variables that are not just predictive, but potentially actionable. For example, identifying which lab values or patient characteristics actually influence recovery, rather than those that merely correlate with it, can lead to better diagnostics, risk stratification, and treatment planning. CausalDRIFT is implemented in an easy-to-use Python package and can be applied to both continuous and categorical features. It provides an interpretable, robust, and clinically grounded alternative to traditional feature selection, bringing causal reasoning into the core of predictive modeling for healthcare.

## 2 Related Work

The challenge of identifying relevant and interpretable features in high-dimensional medical datasets has inspired a diverse body of research across traditional statistics, machine learning, and causal inference. Classical feature selection methods aim to reduce dimensionality and improve model efficiency, while newer approaches seek robustness and causal interpretability. In this section, we review key developments in feature selection, with a focus on their application to medical data, and discuss recent advances in causal reasoning that motivate our proposed method, CausalDRIFT.

### Traditional Feature Selection and Dimensionality Reduction

Feature selection and dimensionality reduction are longstanding techniques in machine learning for mitigating the curse of dimensionality, reducing model complexity, and improving generalization. In the context of healthcare, these methods are often employed to handle high-dimensional clinical data such as lab measurements, physiological signals, and genomic profiles.

### Unsupervised Dimensionality Reduction

Principal Component Analysis (PCA) [17] and Independent Component Analysis (ICA) [3] are widely used unsupervised methods. PCA transforms the original features into a set of orthogonal components that capture the maximum variance, while ICA seeks statistically independent latent sources. Though effective at reducing redundancy, these methods are blind to the outcome variable and can retain components that are irrelevant or even misleading for prediction tasks. Furthermore, the resulting transformed features are typically linear combinations of all original variables, which complicates interpretability in clinical settings.

### Supervised and Embedded Methods

Regularized regression models such as LASSO and Elastic Net [9] perform feature selection as part of the model fitting process. These methods penalize regression coefficients to induce sparsity and are widely used in clinical risk modeling. While outcome-aware, they rely on correlational associations and are sensitive to multicollinearity and confounding bias.

### Filter and Wrapper Approaches

Simple filter methods, such as ranking features based on correlation coefficients or mutual information, are computationally efficient but univariate, failing to account for interactions and joint dependencies. Wrapper-based techniques like Recursive Feature Elimination (RFE) [19] use a predictive model to iteratively prune less important features. Although more accurate, they are computationally expensive and also rely on statistical associations rather than causal relationships.

### Feature Selection in Medical Machine Learning

Feature selection remains pivotal in medical machine learning, where data routinely span demographics, laboratory panels, imaging-derived phenotypes, and longitudinal treatments. Compact feature sets not only improve statistical efficiency but also determine whether a model’s output can be trusted in the clinic.

### Embedded and hybrid approaches

Penalized linear models (e.g., LASSO, Elastic Net) and boosted tree ensembles continue to dominate day-to-day clinical modelling because they embed feature selection in the training objective. A recent example is *DeepSelective*, which augments gradient-gated sparsity with an autoencoder compression stage to yield clinician-verifiable prognostic signatures from large EHRs [20]. Such hybrids illustrate an accelerating trend: coupling sparsity with representation learning to keep interpretability while squeezing performance from high-dimensional records.

### Unsupervised reduction

Principal-component analysis and stacked autoencoders are still widely used to tame omics and imaging volumes; however, a 2024 scoping review of hepatocellular-carcinoma studies shows that deep autoencoders increasingly replace hand-engineered reduction pipelines, yet still provide little transparency on clinical relevance [21].

### Post-hoc explainability

Model-agnostic tools such as SHAP and gradient-based saliency remain popular, but evidence is mounting that explanation techniques should be integrated earlier. For instance, a Scientific Reports study combines Grad-CAM heat-maps with feature-ranking to drive clinician feedback loops in brain-tumour diagnosis, demonstrating how explainability can sharpen—not merely interpret—feature selection [22].

### Distributional robustness

Multi-institution deployment often breaks models whose features encode site-specific shortcuts. A multi-hospital diabetes-readmission study introduced ERR-DGN, showing that explicitly regularising features for domain generalisation outperforms site-specific tuning [23]. Nevertheless, large-scale empirical work from NeurIPS 2024 cautions that “causal” feature subsets do not automatically generalise better than purely predictive ones, underscoring the need for principled selection beyond intuition [24].

### Toward causal relevance

The community is therefore pivoting toward techniques that score variables by estimated causal effects. Recent advances include invariant causal-prediction extensions to mixed outcomes [25], and our own CausalDRIFT, which ranks covariates via residualised treatment effects to produce portable, clinically meaningful feature panels.

### Causality in Feature Selection

In recent years, there has been a growing interest in incorporating causal reasoning into feature selection to address the limitations of purely correlational methods. Traditional approaches often fail to distinguish between features that are causally influential and those that are merely associated with the outcome due to confounding, mediation, or reverse causation. This distinction is especially critical in healthcare, where decisions based on spurious associations can lead to ineffective or harmful interventions. Causal feature selection methods aim to identify features that lie on causal paths to the outcome, or those that would change the outcome under hypothetical interventions. Early works in this domain leverage constraint-based algorithms such as PC and FCI [26], which use conditional independence tests to infer the structure of causal graphs and isolate relevant parent or ancestor variables. These methods are theoretically grounded but struggle with reliability in high-dimensional, noisy, or small-sample settings common in real-world data.

More recent developments focus on learning feature sets that are invariant across environments, based on the principle that causal relationships are stable under interventions or distributional shifts. Approaches such as Invariant Risk Minimization (IRM) [27] and Anchor Regression [28] formalize this idea by selecting features whose predictive utility persists across different data-generating regimes. While promising, these methods often require access to multiple environments or domain labels, which may not always be available in practice. Another important strand of work involves model-based causal effect estimation, such as using treatment effect modeling, propensity score matching, or double machine learning (DML) [16]. These approaches provide robust estimates of a feature’s causal impact on an outcome by adjusting for confounding via flexible machine learning models. However, they are typically applied to a small number of prespecified treatments rather than exhaustively screening the entire feature space. In contrast, our proposed method CausalDRIFT applies DML-style residualization iteratively across all features, estimating average treatment effects (ATEs) while adjusting for confounding in a data-driven way. It thus bridges the gap between scalable feature selection and principled causal inference, providing a practical tool for robust and interpretable variable selection in complex, observational healthcare datasets.

Despite the richness of existing approaches, most feature selection methods fall into one of two categories: they are either outcome-blind (as in unsupervised reductions) or correlation-focused (as in supervised selectors), both of which may lead to unstable or misleading models in the presence of confounding. While recent causal feature selection frameworks offer promising alternatives, they often require access to explicit graph structures or environment labels, limiting their practicality in clinical applications. Our work contributes to this emerging space by introducing a causally grounded, model-agnostic, and computationally feasible approach that estimates the effect of each feature on the outcome via principled residualization. CausalDRIFT thus advances the field by aligning feature selection with causal reasoning, enabling more interpretable and robust modeling in healthcare settings.

## 3 Method

### 3.1 CausalDRIFT: Algorithm Description

CausalDRIFT is a causal feature selection algorithm that leverages the Frisch-Waugh-Lovell (FWL) theorem and Double Machine Learning (DML) principles to estimate the average treatment effect (ATE) of each feature on the target outcome, controlling for confounding influences. Unlike correlation-based feature selection methods, CausalDRIFT identifies variables with potential *causal influence*, thereby enhancing interpretability and robustness in downstream predictive models.

The core idea is to estimate the ATE of each candidate feature *T*_*j*_ (treated as a putative treatment) on the outcome *Y*, after adjusting for a data-driven set of confounders *X*. This is achieved through residualization and a final linear regression step.

#### Algorithm 1

CausalDRIFT: Causal Feature Selection via Residualization

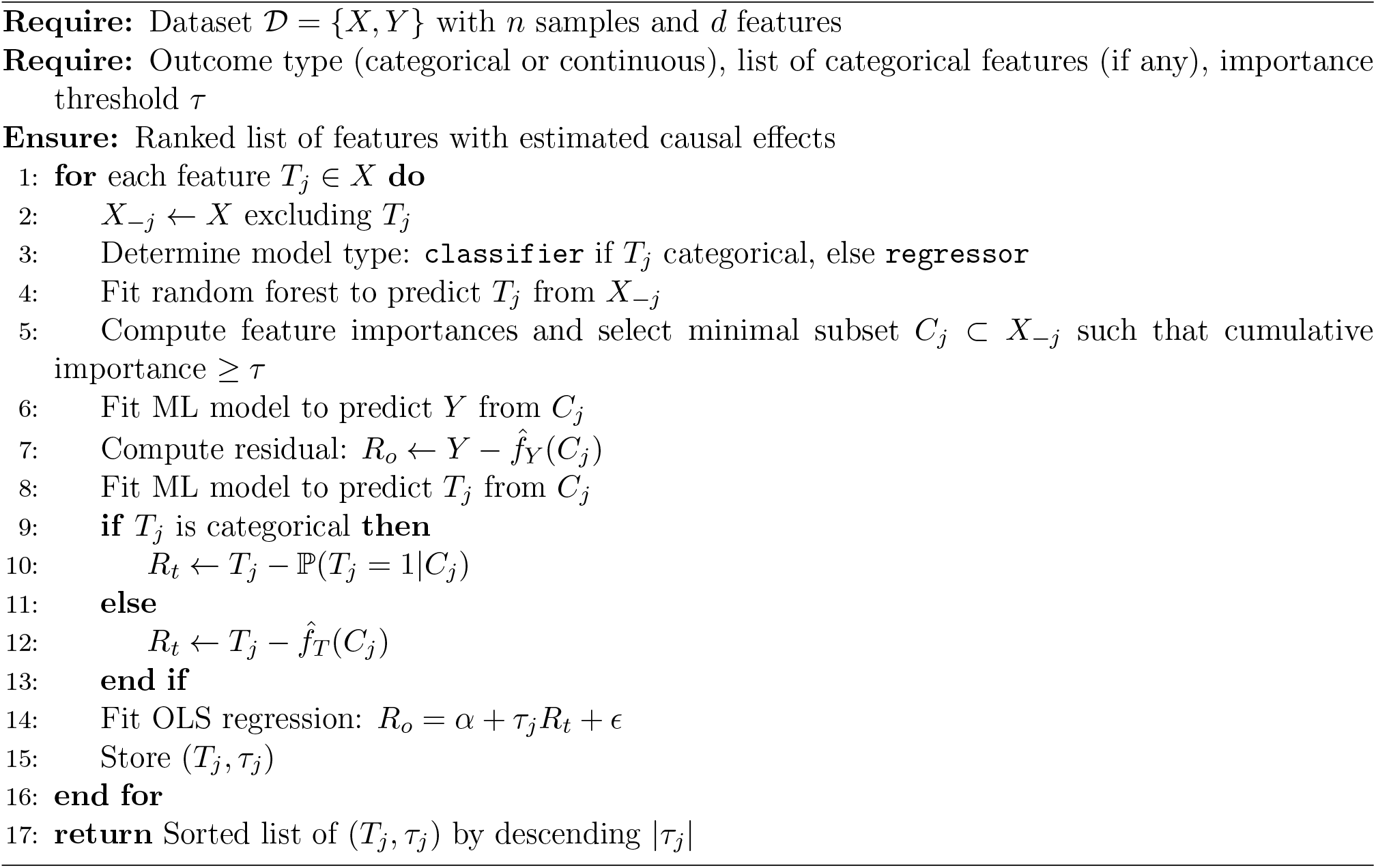

**Step 1: Confounder Selection** For each feature *T*_*j*_, we first identify a subset of variables *X*_*−j*_ that serve as potential confounders. We fit a random forest model to predict *T*_*j*_ from *X*_*−j*_, and retain the smallest subset of features that cumulatively explain a fixed threshold (e.g., 80%) of the total feature importance:

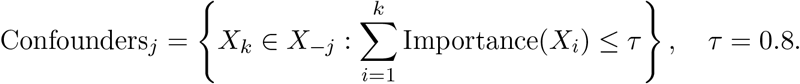

This provides a heuristic for backdoor adjustment while keeping the confounder set compact and interpretable.

**Step 2: Residualization via Machine Learning** Given the selected confounders *C*_*j*_, we perform two residualization steps:

1. Fit a model 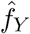 (random forest regressor or classifier) to predict the outcome *Y* from *C*_*j*_. Compute the outcome residual:

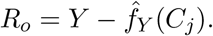
2. Fit a model 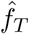 (regressor or classifier depending on the type of *T*_*j*_) to predict *T*_*j*_ from *C*_*j*_. Compute the treatment residual:

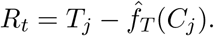

For categorical outcomes or treatments, probabilistic classifiers are used, and the residual is computed with respect to the predicted probability (e.g., for binary treatment, *R*_*t*_ = *T*_*j*_ *−* ℙ (*T*_*j*_ = 1|*C*_*j*_)).

**Step 3: Causal Effect Estimation** We then perform a simple linear regression of *R*_*o*_ on *R*_*t*_:

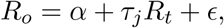

where *τ*_*j*_ is interpreted as the estimated average treatment effect (ATE) of feature *T*_*j*_ on the outcome *Y*. This step is justified by the FWL theorem, which states that in a linear setting, regressing residuals yields the same coefficient as a full multiple regression.

**Figure 1:**
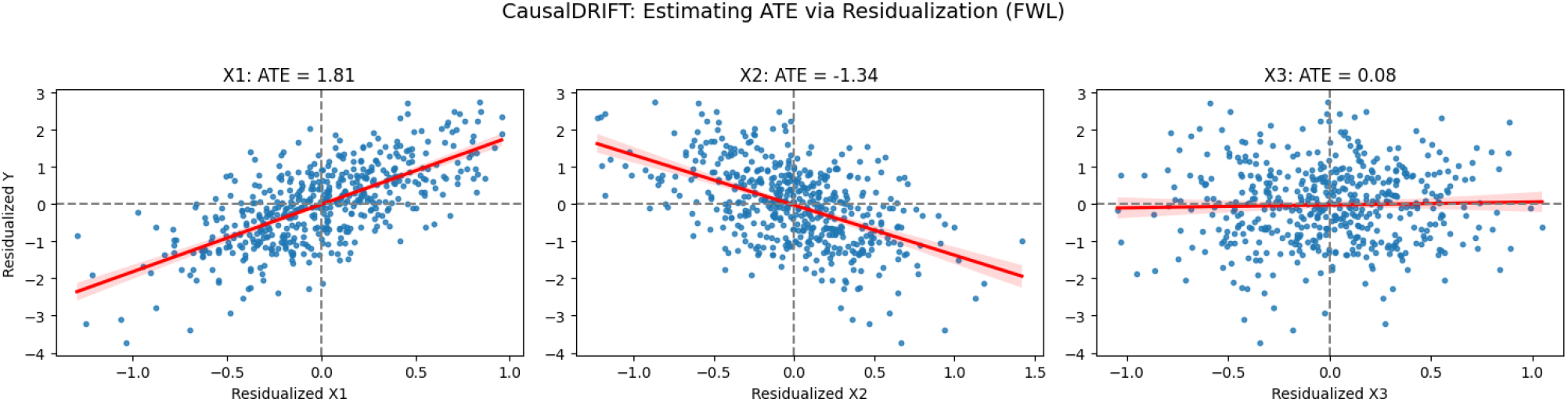
Estimating ATE via Residualization (FWL)

Figure 1 illustrates how CausalDRIFT estimates the Average Treatment Effect (ATE) of each feature using residualization and the Frisch–Waugh–Lovell (FWL) theorem. For each feature, the residualized treatment (*R*_*t*_) is plotted against the residualized outcome (*R*_*o*_), and a linear regression is fitted to estimate the causal effect. As shown, feature **X1** has a strong positive ATE of **1.82**, and **X2** has a strong negative ATE of **-1.34**, indicating their significant causal influence on the outcome. In contrast, **X3**, a non-causal noise feature, shows an ATE close to zero (**0.08**), confirming its irrelevance. This visual supports the effectiveness of CausalDRIFT in isolating causal contributions, even in the presence of confounding variables.

#### Output: Feature Ranking

The algorithm iterates over all features in the dataset and returns a ranked list of features ordered by the magnitude of their estimated causal effects:

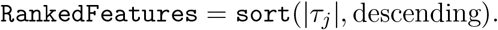

Features with larger |*τ*_*j*_| are considered to have stronger causal influence and are retained for downstream modeling.

#### Handling Categorical Variables

All features are treated numerically, but users may specify a list of features that encode discrete categories (e.g., education level). For these, classification models are used in the residualization steps, and residuals are computed using predicted probabilities, ensuring that the ATE regression remains meaningful even for categorical inputs.

#### Computational Complexity and Practical Relevance

CausalDRIFT is computationally efficient for moderate to large medical datasets. The primary computational cost arises from fitting random forest models for each feature to (i) identify confounders, and (ii) perform residualization of the treatment and outcome. Let *d* be the number of features and *n* the number of samples. Each random forest has a training complexity of approximately *𝒪*(*n* log *n*) per tree, and the number of such models scales linearly with *d*. Therefore, the total time complexity is approximately *𝒪*(*d · n* log *n*), which is tractable given that forests are highly parallelizable.

In terms of space complexity, the algorithm stores residual vectors and subsets of features for each iteration, resulting in 𝒪 (*nd*) space in the worst case. However, since confounders are typically sparse (e.g., top 80% by importance), the effective memory usage remains low for most real-world use cases. This efficiency is particularly advantageous in clinical applications where datasets often involve hundreds of variables but limited sample sizes, such as intensive care records, lab panels, or patient risk factors. By avoiding full pairwise conditioning and instead using greedy heuristics for confounder selection, CausalDRIFT balances statistical rigor with scalability. The ability to retain causally relevant and interpretable features without extensive preprocessing makes it highly suitable for medical environments where both transparency and computational efficiency are essential.

#### Strengths of the Algorithm

CausalDRIFT offers several strengths that make it particularly well-suited for high-stakes domains like healthcare. First, it grounds feature selection in causal theory, ensuring that retained variables reflect plausible cause-effect relationships rather than spurious correlations. This leads to more robust and clinically interpretable models, which are essential for real-world medical deployment. Second, the algorithm naturally accommodates non-linearities and high-dimensional interactions by employing machine learning models, such as random forests for residualization, extending the classical Frisch-Waugh-Lovell (FWL) approach beyond linear settings. Third, it provides an intuitive and quantitative measure of causal influence (the average treatment effect) for each feature, enabling not just selection, but also prioritization of variables for clinical insight or intervention design. Finally, its modular design allows easy adaptation: researchers can swap in alternative learners (e.g., gradient boosting, neural nets) for the residualization steps, or refine confounder selection strategies based on domain knowledge, making CausalDRIFT a flexible foundation for causal feature discovery.

### 3.2 Baseline Methods

To evaluate the performance of CausalDRIFT, we primarily compare it against seven widely used feature selection and dimensionality reduction methods, each representing a distinct methodological category: unsupervised projection, statistical association, regularized regression, and wrapper-based approaches. All methods are applied prior to training the downstream predictive model, and the number of retained features is standardized for fair comparison. For the first primary datasets, we assesed our algorithm’s performance with Principle Component Analysis, Independent Component Analysis, Correlation Coefficient, Elastic Net, and Recursive Feature Elimination. For the final complex dataset, we additionally assesed our algorithm with Mutual Information, Autoencoder, and Tree-based (XGBoost) feature selection.

#### Principal Component Analysis (PCA)

PCA is a linear dimensionality reduction technique that transforms the original feature space into a set of orthogonal components that maximize variance [17]. Given a data matrix *X ∈* ℝ^*n×d*^, PCA computes a decomposition *X* = *U* Σ*V*^*T*^ via singular value decomposition (SVD), and retains the top-*k* principal components that explain the majority of variance in the data. Although widely used, PCA is agnostic to the outcome variable and does not account for causal relationships.

#### Independent Component Analysis (ICA)

ICA seeks a linear transformation of the data such that the resulting components are statistically independent [3]. Unlike PCA, which captures decorrelation, ICA focuses on higher-order independence. This can be effective when the underlying data-generating process involves latent independent sources. However, ICA similarly disregards any relationship to the outcome and may select components that are statistically independent but causally irrelevant.

#### Elastic Net

Elastic Net is a regularized regression model that combines L1 (lasso) and L2 (ridge) penalties to encourage both sparsity and stability in feature selection [9]. It solves the following optimization problem:

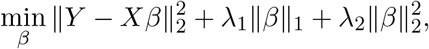

where *λ*_1_ and *λ*_2_ control the sparsity and shrinkage, respectively. Elastic Net is supervised and outcome-aware, but still purely associational in its selection criteria.

#### Autoencoder

Autoencoders are unsupervised neural networks that learn a compressed representation (encoding) of the input data by minimizing reconstruction error [18]. A typical autoencoder consists of an encoder *f*_*θ*_ : ℝ^*d*^ *→* ℝ^*k*^ and decoder *g*_*ϕ*_ : ℝ^*k*^ *→* ℝ^*d*^ trained jointly to minimize:

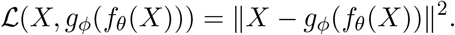

While powerful for dimensionality reduction, autoencoders do not use the outcome *Y* during training, and the learned latent features are typically not interpretable.

#### Correlation Coefficient

This is a simple filter-based method that selects features based on their univariate correlation with the outcome variable. For each feature *X*_*j*_, the Pearson correlation coefficient with the outcome *Y* is computed as:

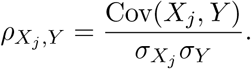

Features with the highest |*ρ*| values are retained. While easy to compute, this method is highly sensitive to confounding and spurious associations.

#### Recursive Feature Elimination (RFE)

RFE is a wrapper-based feature selection method that recursively eliminates the least important features based on a model’s learned weights [19]. In each iteration, a model (e.g., linear regression or tree-based) is trained, features are ranked by importance, and the lowest-ranked feature is removed. This continues until a desired number of features remains. RFE is supervised and model-aware but computationally expensive and still vulnerable to including non-causal but predictive variables.

Together, these baselines represent a comprehensive set of strategies commonly used in healthcare machine learning pipelines. However, none of them explicitly account for causal relationships between features and outcomes, which motivates the need for methods such as CausalDRIFT.

#### Mutual Information (MI) Feature Selection

Mutual Information (MI) measures the amount of shared information between a feature *X*_*i*_ and the target variable *Y*. It quantifies how much knowing *X*_*i*_ reduces the uncertainty of *Y*. The mutual information between a discrete feature *X*_*i*_ and the target *Y* is defined as:

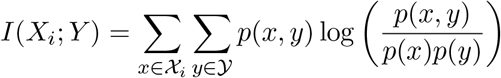

where *p*(*x, y*) is the joint probability distribution of *X*_*i*_ and *Y*, and *p*(*x*), *p*(*y*) are the marginal distributions. Features with higher mutual information scores are considered more relevant for predicting the target.

#### Tree-Based Feature Selection (XGBoost)

XGBoost is a gradient-boosted decision tree algorithm that provides built-in feature importance scores. During training, it evaluates how often and how effectively each feature is used to split data across all trees in the ensemble.

Feature importance in XGBoost can be measured using:

- **Gain**: The average reduction in loss (e.g., log loss or mean squared error) when a feature is used for splitting.
- **Frequency (Weight)**: The number of times a feature is used in all trees.
- **Cover**: The number of data samples affected by splits on a feature. Mathematically, the gain from a split using feature *X*_*i*_ is:

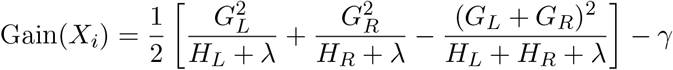

where *G*_*L*_, *G*_*R*_ and *H*_*L*_, *H*_*R*_ are the gradients and Hessians for the left and right child nodes respectively, and *λ, γ* are regularization parameters. XGBoost ranks features by aggregating these metrics across all trees to identify the most influential predictors.

## 4 Experiments

We designed a comparative evaluation to assess the effectiveness of CausalDRIFT relative to widely used feature selection methods across multiple healthcare prediction tasks. Our experimental setup was designed to isolate the contribution of feature selection by holding the downstream predictive model and evaluation metrics constant across all conditions. Reproducibility guidelines are accessible from the GitHub repository of this research: https://github.com/SakibHasanSimanto/causalsoap

### 4.1 Datasets

We selected four publicly available clinical datasets from Kaggle, covering diverse medical domains:

- **Cleveland Heart Disease Dataset:** Includes 13 features such as chest pain type, cholesterol, and resting ECG to predict the presence of heart disease.
- **Breast Cancer Wisconsin Dataset:** Comprises 31 numeric features derived from digitized images of breast mass biopsies, used to classify tumors as benign or malignant.
- **Pima Indians Diabetes Dataset:** Contains 8 physiological variables (e.g., BMI, glucose level, blood pressure) to predict diabetes onset.
- **Polysystic ovary syndrome (PCOS) Dataset**: Contains 44 variables to predict PCOS.

All datasets were clean and free from missing values, and therefore required no additional preprocessing. The target variable in each dataset is binary, representing the presence or absence of a medical condition. Among the datasets, Cleveland Heart Disease Dataset has some categorical features that helps in stress testing CausalDRIFT. To measure causality, we specially assessed our algorithm on PCOS dataset, since the cause of this disease is unknown.

### 4.2 Experimental Pipeline

Each dataset (except PCOS) was processed using the following pipeline:

1. **Feature Selection:** We applied six feature selection or dimensionality reduction methods: Principal Component Analysis (PCA), Independent Component Analysis (ICA), Correlation Coefficient (CorrCoef), Recursive Feature Elimination (RFE), Elastic Net, and CausalDRIFT. These methods reduced the number of features by 3 times, as shown in Table 1.
2. **Dataset Generation:** For each method, a transformed or reduced version of the dataset was created, yielding a total of 18 datasets (3 original datasets × 6 selection methods).
3. **Model Training:** An XGBoost classifier was trained on each of the 18 datasets using default hyperparameters for unbiased observation.
4. **Evaluation Metrics:** We evaluated model performance using four standard classification metrics: Accuracy, Precision, Recall, and F1-Score.
5. **Causality and Complexity Test:** To test CausalDRIFT’s performance on complex dataset, we compared the method with three other feature selection methods: Mutual Information, Autoencoder, Tree-based (XGBoost) feature selection.

**Table 1:**
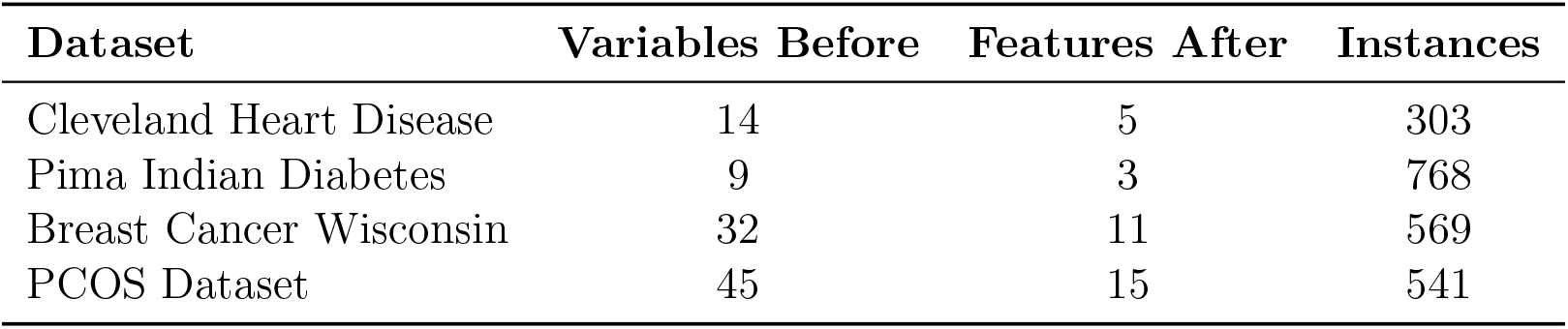
Dataset dimensions before and after feature selection (approximately reducing features by a factor of 3)

| Dataset | Variables Before | Features After | Instances |
| --- | --- | --- | --- |
| Cleveland Heart Disease | 14 | 5 | 303 |
| Pima Indian Diabetes | 9 | 3 | 768 |
| Breast Cancer Wisconsin | 32 | 11 | 569 |
| PCOS Dataset | 45 | 15 | 541 |

Feature selection techniques were applied to reduce the number of input features by approximately a factor of 3. For example, the Cleveland Heart Disease dataset initially had 14 variables (13 features and 1 outcome), which after division by 3 gives 14 *÷* 3 = 4.67, rounded to 5 selected features.

By applying all six feature selection methods to each dataset independently, we were able to analyze the comparative influence of feature selection on downstream predictive performance. The use of XGBoost as a consistent model choice allowed us to focus purely on the effect of selected features, rather than variations in model architecture or optimization. We recorded accuracy, precision, recall, and f1-score for all datasets, and aggregated the results across datasets. This provided both per-dataset and cross-dataset insights into which methods offered the most robust and accurate feature sets under consistent modeling conditions. This experimental design enables us to directly compare how well CausalDRIFT performs in extracting causally meaningful and predictively strong features relative to conventional selection techniques. The results, discussed in the next section, highlight both the predictive performance and interpretability advantages of our approach across diverse clinical datasets.

### 4.3 Statistical Significance Testing

To assess whether differences in model performance across feature selection methods are statistically significant, we conducted a controlled significance test using the Cleveland Heart Disease dataset. Since Cleveland dataset contains both categorical and continuous features, it will help stress testing our algorithm. We focused on the recall score as the evaluation metric, since in clinical applications, false negatives—i.e., failing to detect a true condition can be far more detrimental than false positives.

For each of the six feature selection methods (PCA, ICA, Correlation Coefficient, RFE, Elastic Net, and CausalDRIFT), we trained and evaluated an XGBoost classifier 30 independent times. Running the experiment 30 times leverages the Central Limit Theorem, which states that the sampling distribution of the mean will approximate a normal distribution as the number of samples increases, typically becoming reliable around n ≥ 30. This makes parametric tests (ANOVA in our case) statistically valid, as they assume normally distributed sample means [29].

**Figure 2:**
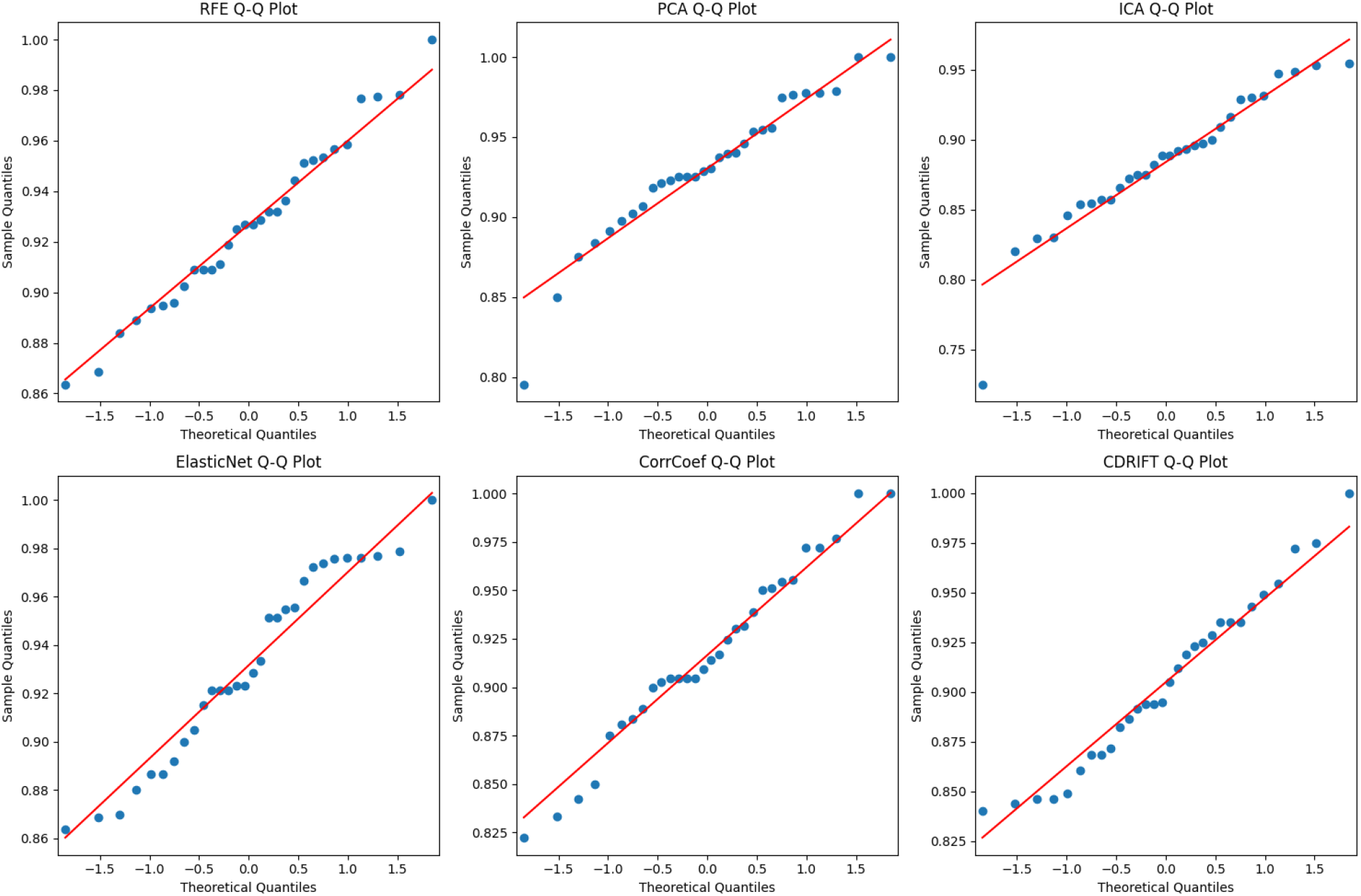
Q-Q plots of recall scores for different methods

Q–Q plots in Figure 2 compare the quantiles of the observed data to a standard normal distribution. If the points lie along the red diagonal, the data are approximately normally distributed, which is a key assumption of ANOVA. CDRIFT, CorrCoef, RFE, PCA: The recall scores follow the red line closely, meaning that the normality assumption is well met. ElasticNet is mostly normal, with a slight upward bend in the tail, suggesting mild right-skew. ICA deviates more strongly from the line, especially in the lower quantiles. It suggests deviation from normality, possibly due to outliers or a skewed distribution.

So, 30 iterations provide a strong foundation for inference and significance testing, even if the underlying recall scores are not perfectly normal. In each run, we omitted the use of a fixed random_state parameter in data splitting or model training, thereby introducing randomness in both training and test partitions. This procedure generates a distribution of recall scores for each method, providing a robust estimate of model performance under varying data conditions. With 30 recall scores collected per method, we tested for significant differences in the mean recall across all feature selection methods using statistical hypothesis testing. Our objective was to determine whether CausalDRIFT yields a statistically higher recall compared to traditional techniques. This multi-run evaluation framework allows us to account for stochastic variation in model fitting and strengthens the reliability of the observed performance trends.

## 5 Results

We evaluate our proposed causal inference-based feature selection method, CausalDRIFT, against established feature selection and dimensionality reduction techniques on several dataset. All experiments were conducted using the XGBoost classifier to ensure consistency in the classification model. The primary aim is to assess whether causal reasoning in feature selection can provide performance benefits or interpretability advantages compared to conventional methods.

### 5.1 Evaluation Metrics

The evaluation metrics used in this study include:

- **Accuracy**: Proportion of total correct predictions.
- **Precision**: Proportion of true positives among predicted positives.
- **Recall**: Proportion of true positives among actual positives.
- **F1-Score**: Harmonic mean of precision and recall.

### 5.2 Results Summary on Heart Disease Dataset

Figure 3 visualizes the classification performance of XGBoost when trained on features selected by six different methods.

**Figure 3:**
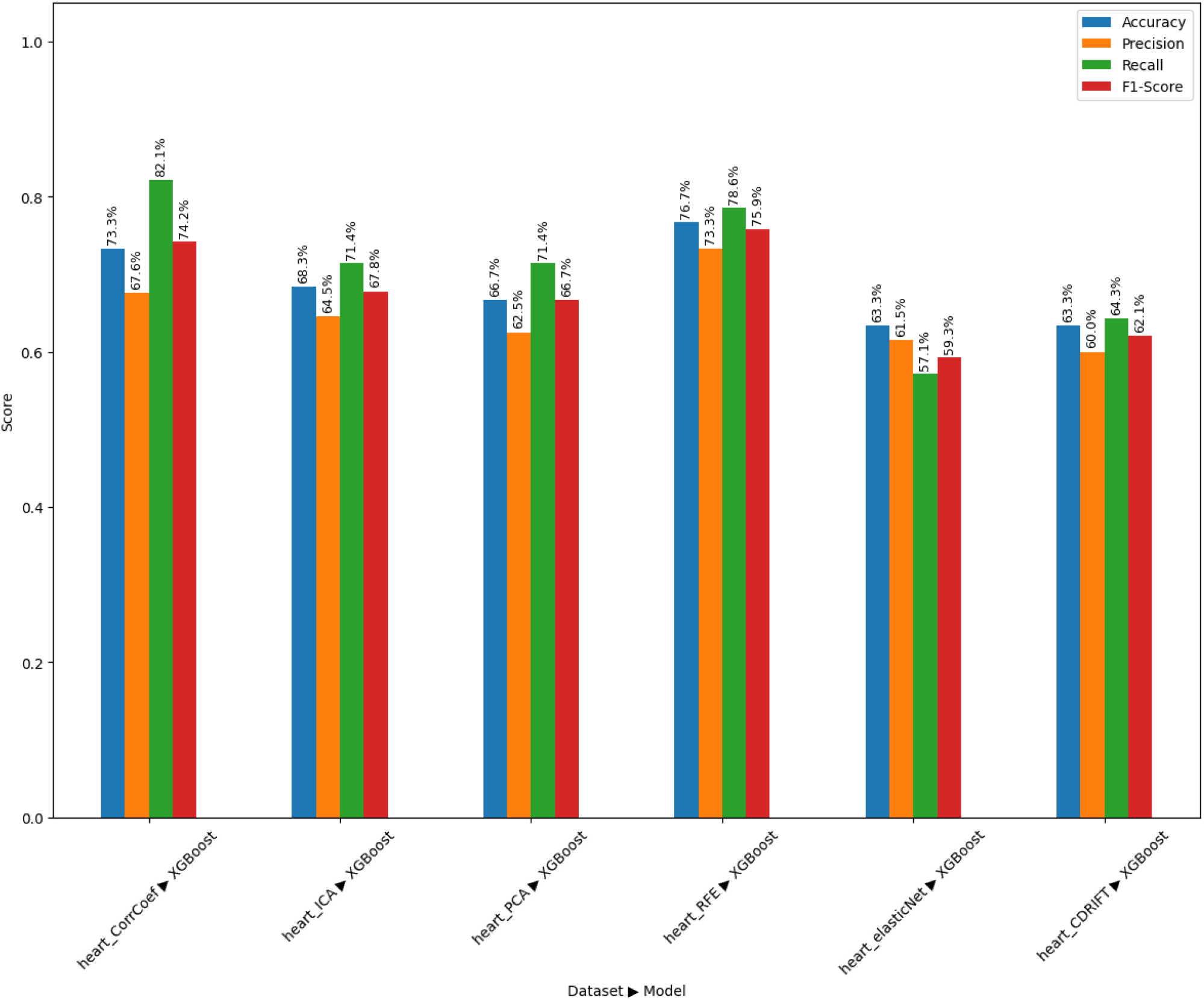
Performance Comparison of Feature Selection Methods (Heart Disease Dataset)

The experimental results indicate that CausalDRIFT achieves an accuracy of 63.3%, which is competitive with ElasticNet and PCA, although it lags behind CorrCoef, ICA, and RFE. In terms of F1-Score, CausalDRIFT (0.621) outperforms ElasticNet (0.593), but falls short of the other methods. The strongest performing method is RFE, achieving the highest accuracy (76.7%) and F1-Score (75.9%), followed by CorrCoef, which exhibits the highest recall (82.1%). Notably, while CausalDRIFT shows moderate recall (64.3%) and precision (60.0%), this balance suggests the method selects causally relevant features rather than those merely correlated with the target. Although CausalDRIFT does not surpass the benchmark methods in raw predictive performance, its strength lies in offering a causally interpretable and potentially more generalizable feature subset. This characteristic is essential in sensitive domains such as healthcare, where understanding *why* a model makes a decision is as important as the decision itself. The results demonstrate that while CausalDRIFT currently underperforms compared to traditional methods in terms of accuracy and F1-Score, it provides a valuable trade-off between interpretability and performance.

### 5.3 Results Summary on Diabetes Dataset

Figure 4 shows the comparative results of the feature selection methods on the diabetes dataset.

**Figure 4:**
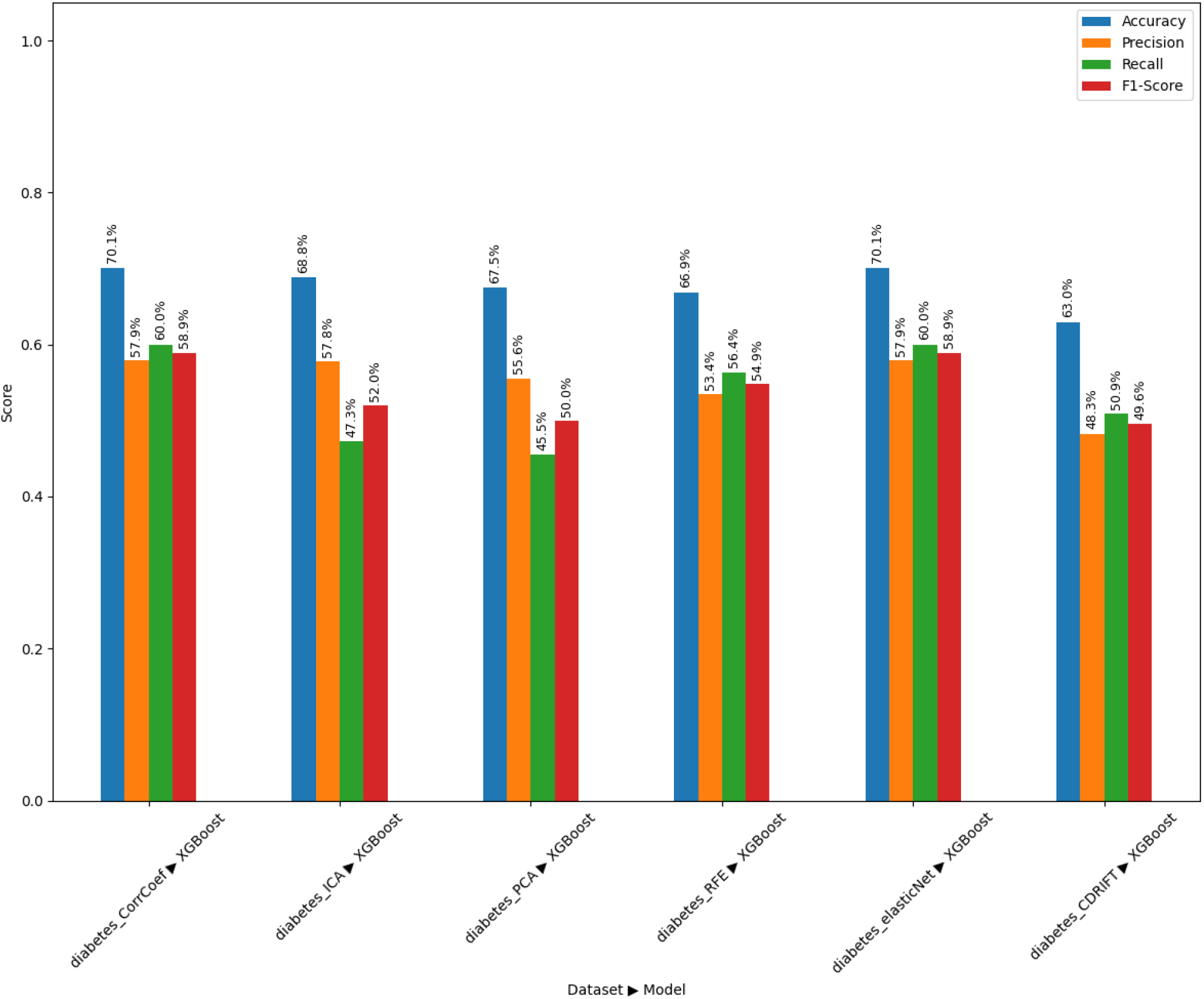
Performance Comparison of Feature Selection Methods (Diabetes Dataset)

On the diabetes dataset, CorrCoef and ElasticNet achieved the best overall performance, each recording an accuracy of 70.1% and an F1-score of 58.9%. Among traditional methods, PCA and ICA under-performed slightly in both recall and F1-score. CausalDRIFT achieved an accuracy of 63.0% and an F1-score of 49.6%, which is lower than all baseline methods. While its precision and recall are relatively balanced, the overall scores suggest that the features selected by CausalDRIFT, though causally meaningful, might be insufficiently predictive on this dataset without further refinement. The results indicate that for the diabetes dataset, conventional correlation- or regularization-based feature selection methods may currently outperform causality-based approaches in pure predictive terms. However, this may reflect the complex and less causally structured nature of the diabetes dataset, where statistical signal may dominate over causal relationships in determining outcomes. Even though CausalDRIFT underperforms compared to other methods on the diabetes dataset in terms of predictive metrics, its potential lies in interpretability and robustness to spurious correlations.

### 5.4 Results Summary on Breast Cancer Dataset

Figure 5 summarizes the classification performance of each method on the breast cancer dataset.

**Figure 5:**
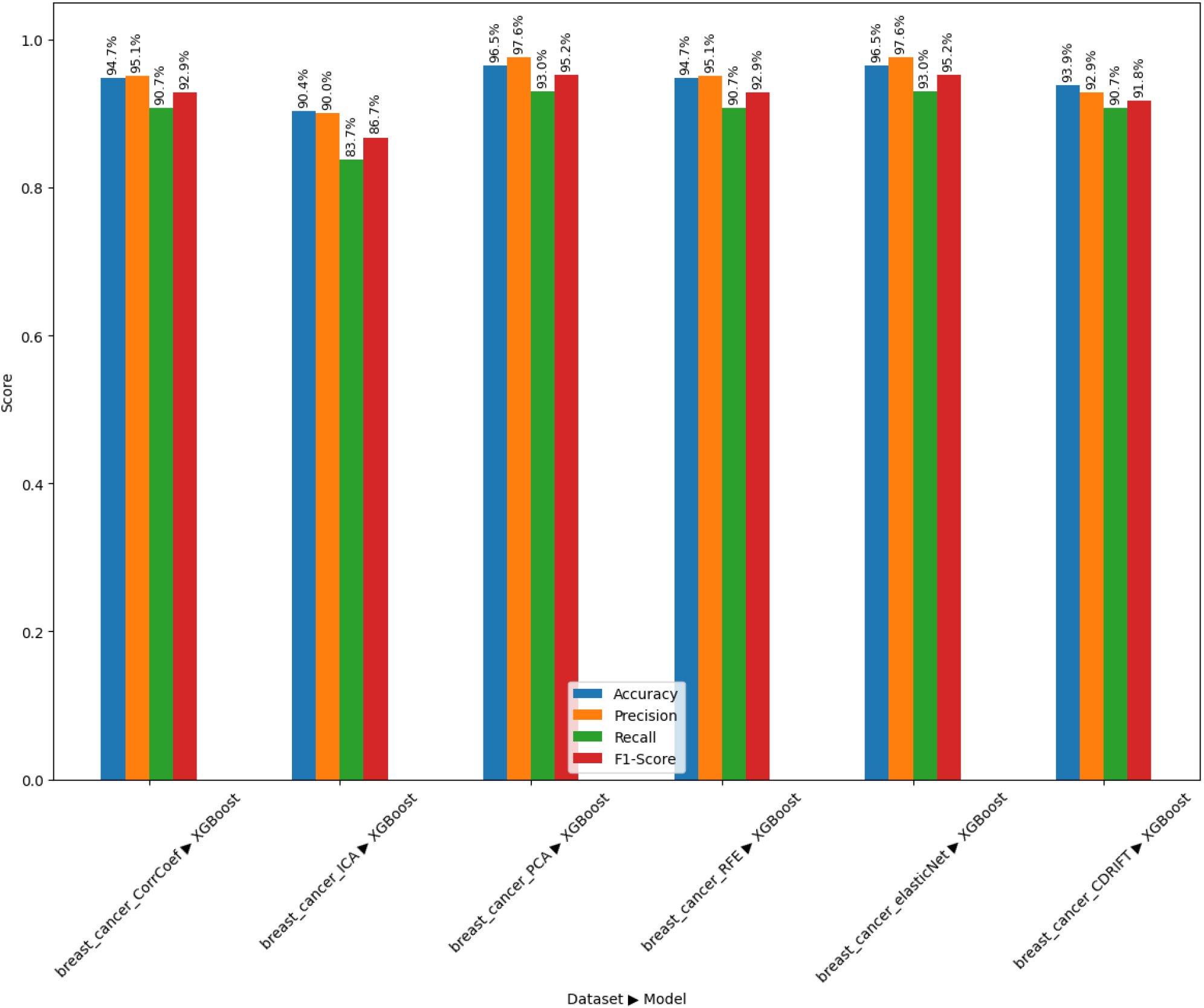
Performance Comparison of Feature Selection Methods (Breast Cancer Dataset)

The results show that all methods performed well on the breast cancer dataset, with PCA and ElasticNet achieving the highest scores across all metrics (accuracy: 96.5%, F1-score: 95.2%). CausalDRIFT achieved an accuracy of 93.9% and an F1-score of 91.8%, which is slightly lower than the top-performing models but still competitive. Notably, its recall (90.7%) matches that of CorrCoef and RFE, indicating strong sensitivity. This suggests that CausalDRIFT captures most of the relevant causal factors linked to positive diagnoses while maintaining solid predictive performance. The bar heights of CausalDRIFT also indicates its performance consistency accorss the metrics, that the other algorithms could not achieve.

**Table 2:** Standard Deviation of Performance Metrics in Breast Cancer Dataset.

| Feature Selection Method | Standard Deviation |
| --- | --- |
| Correlation Coefficient | 1.74 |
| Independent Component Analysis | 2.72 |
| Principal Component Analysis | 1.71 |
| Recursive Feature Elimination | 1.74 |
| Elastic Net | 1.71 |
| **CausalDRIFT** | **1.19** |

Table 2 shows that the CausalDRIFT algorithm has relatively lower fluctuations in the performance metrics (accuracy, precision, recall, f1-score), which demonstrates its performance consistency in the dataset compared to the other methods.

While the precision of CausalDRIFT (92.9%) is slightly lower than PCA and ElasticNet (97.6%), it still demonstrates robustness, especially given its focus on causal rather than purely correlational signal. On the breast cancer dataset, CausalDRIFT performs comparably with state-of-the-art methods, demonstrating that causally selected features can achieve near-optimal and consistent predictive performance. The method’s success on this dataset supports its potential utility in clinical applications where both accuracy and interpretability are essential. These results reinforce the viability of CausalDRIFT as a general-purpose causal feature selection strategy with minimal compromise on predictive quality, and relatively higher consistency across performance metrics.

**Table 3:** CausalDRIFT Performance Summary Across Datasets.

| Dataset | Causal Structure | Feature Type | CausalDRIFT Performance |
| --- | --- | --- | --- |
| Heart Disease | Medium | Mixed (numeric + categorical) | Moderate |
| Breast Cancer | Strong | All numeric | Strong |
| Diabetes | Weak/latent | All numeric | Weak |

Table 3 summarizes that CausalDRIFT is most effective in datasets where causal relationships are strong, and dataset initially has higher number of features. Its performance diminishes in the presence of latent confounders or when predictive power stems primarily from statistical correlation rather than causal influence (e.g., Diabetes). Despite occasional trade-offs in raw performance, CausalDRIFT offers the added value of interpretability and robustness with minimal performance loss, which are critical in clinical and decision-sensitive applications.

### 5.5. Performance Evaluation on PCOS Dataset

For a robust and final evaluation, we tested our algorithm on PCOS dataset, a disease whose causality is yet to be found. Polycystic Ovary Syndrome (PCOS) is a complex endocrine disorder with multifactorial and not yet fully understood etiology. To evaluate the applicability of our proposed causal inference-based feature selection algorithm CausalDRIFT, we applied it to the PCOS dataset, aiming to identify influential features even when the underlying cause of the disease is unknown. The selected features were used to train an XGBoost classifier, and the performance was compared against several additional baseline feature selection and dimensionality reduction techniques, including Mutual Information, Autoencoder, and tree-based importance from XGBoost.

Table 4 presents the classification performance metrics (precision, recall, and F1-score) for all methods. Our approach, CausalDRIFT, achieved an accuracy of 90%, with a macro-averaged F1-score of 0.90, outperforming PCA (86%), ICA (90%), and Autoencoder (72%), and matching the performance of tree-based methods and Mutual Information (both at 90%–92%). Notably, CausalDRIFT provided a strong and consistent balance between classes: high precision for class 0 (non-PCOS) and high recall for class 1 (PCOS), indicating effective treatment-outcome disentanglement. Figure 6 illustrates accuracy score of other feature selection methods against CausalDRIFT.

**Table 4:** XGBoost classification results on PCOS dataset using various feature selection methods.

| Method | Accuracy | Macro F1-score | Weighted F1-score |
| --- | --- | --- | --- |
| PCA | 0.86 | 0.86 | 0.86 |
| Mutual Information | **0.92** | **0.92** | **0.92** |
| Elastic Net | 0.89 | 0.89 | 0.89 |
| Correlation Coefficient | 0.87 | 0.87 | 0.87 |
| ICA | 0.90 | 0.89 | 0.90 |
| Tree-based (XGBoost) | 0.90 | 0.90 | 0.90 |
| Autoencoder | 0.72 | 0.72 | 0.72 |
| **CausalDRIFT (Ours)** | **0.90** | **0.90** | **0.90** |

These results validate the robustness of CausalDRIFT in settings where the causal structure is ambiguous or unknown, highlighting its potential as a principled alternative to conventional correlation-based or black-box feature selection techniques.

**Figure 6:**
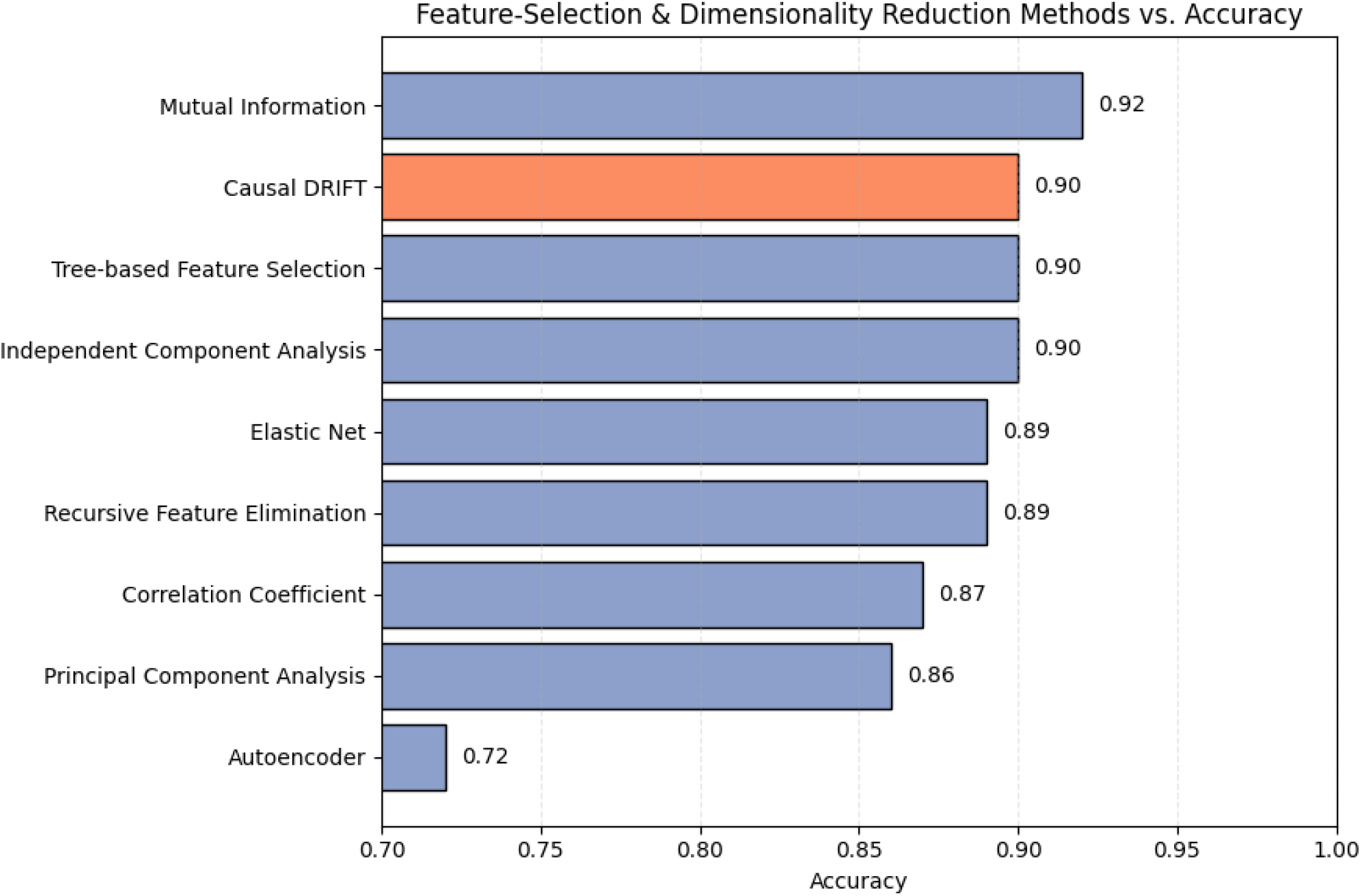
Feature Selection methods vs Accuracy in PCOS Dataset

Interestingly, while CausalDRIFT demonstrated strong performance on datasets such as PCOS and Breast Cancer, where the underlying causal structure is complex or partially unknown, it underperformed on datasets like Diabetes and Heart Disease, which exhibit strong feature-outcome correlations. This suggests that in correlation-dominated datasets, traditional feature selection methods that directly exploit statistical associations may outperform causal approaches. In contrast, CausalDRIFT excels in uncovering hidden causal signals, especially when spurious correlations or latent confounding obscure the true relationships. Thus, its strength lies in scenarios where causal reasoning provides an advantage over purely associative models.

It is also observed that CausalDRIFT performs relatively better on high-dimensional, low-sample size (HDLSS) datasets, such as the Breast Cancer dataset (569 × 32) and the PCOS dataset (541×45), compared to more balanced datasets like the Cleveland Heart Disease (303 × 14) and the Pima Indian Diabetes dataset (768 × 9). This trend may be attributed to the internal causal structure of CausalDRIFT, which prioritizes features based on their average treatment effect (ATE) and potential influence in simulated interventional settings. In high-dimensional, low-sample datasets, conventional correlation-based methods often suffer from overfitting or misleading spurious associations. However, CausalDRIFT leverages the principle of causality, aiming to identify features that have a stable, robust causal influence on the outcome variable, even under data scarcity. Additionally, datasets with more features tend to have a richer set of potential confounding variables and indirect pathways, which CausalDRIFT can exploit to detect truly influential causal factors. This advantage becomes less prominent in datasets with fewer variables, where the causal structure is simpler or nearly saturated, leaving limited room for causal disentanglement. These results suggest that CausalDRIFT may be particularly useful in biomedical settings with high-dimensional feature spaces and limited labeled data, scenarios where traditional statistical feature selection approaches tend to underperform.

### 5.6 ANOVA-Based Comparison of Recall Performance

To formally assess whether differences in recall scores across feature selection methods are statistically significant, we conducted a one-way ANOVA test on the 30 recall values obtained per method using the Cleveland Heart Disease dataset. The test compares the within-group and between-group variances across the six methods.

**Table 5:** One-way ANOVA results comparing recall scores across feature selection methods.

| Source | Sum Sq | df | F-value | p-value |
| --- | --- | --- | --- | --- |
| Method | 0.051343 | 5 | 5.63 | **0.000078** |
| Residual | 0.317515 | 174 | — | — |

The results indicate a statistically significant effect of the feature selection method on recall scores (F(5, 174) = 5.63, p < 0.001). This implies that at least one method yields a mean recall that differs significantly from the others. Given the critical importance of recall in medical diagnostics–where false negatives can be life-threatening–this finding underscores the practical relevance of choosing an effective feature selection method. We also conducted Post hoc analysis (Tukey’s HSD) to identify which specific pairs of methods differ significantly.

### 5.7 Post Hoc Analysis: Tukey HSD Test

To identify which feature selection methods differed significantly in recall performance, we conducted a Tukey Honest Significant Difference (HSD) test following the significant ANOVA result. The test controls the family-wise error rate at *α* = 0.05 and compares all pairs of group means.

**Table 6:** Tukey HSD pairwise comparison of recall means across feature selection methods.

| Group 1 | Group 2 | Mean Diff | p-adj | Lower | Upper | Significant |
| --- | --- | --- | --- | --- | --- | --- |
| CausalDRIFT | CorrCoef | 0.0117 | 0.8977 | -0.0201 | 0.0434 | No |
| CausalDRIFT | ElasticNet | 0.0267 | 0.1562 | -0.0051 | 0.0584 | No |
| CausalDRIFT | ICA | -0.0209 | 0.4106 | -0.0527 | 0.0109 | No |
| CausalDRIFT | PCA | 0.0255 | 0.1954 | -0.0063 | 0.0573 | No |
| CausalDRIFT | RFE | 0.0219 | 0.3526 | -0.0098 | 0.0537 | No |
| CorrCoef | ICA | -0.0325 | 0.0416 | -0.0643 | -0.0007 | **Yes** |
| ElasticNet | ICA | -0.0475 | 0.0004 | -0.0793 | -0.0157 | **Yes** |
| ICA | PCA | 0.0464 | 0.0006 | 0.0146 | 0.0781 | **Yes** |
| ICA | RFE | 0.0428 | 0.0020 | 0.0110 | 0.0746 | **Yes** |

The results show that CausalDRIFT did not differ significantly from any other method in terms of mean recall on the Cleveland dataset (all p > 0.15). However, several pairs involving ICA showed statistically significant differences:

- ICA underperformed relative to ElasticNet (*p* = 0.0004) and CorrCoef (*p* = 0.0416)
- ICA also showed significantly lower recall than PCA and RFE

These findings suggest that while CausalDRIFT is statistically comparable to the strongest traditional methods in terms of recall, some unsupervised techniques like ICA may lead to significantly degraded performance in clinical settings. This reinforces the need for more informed and robust selection strategies, particularly those grounded in causal reasoning.

**Figure 7:**
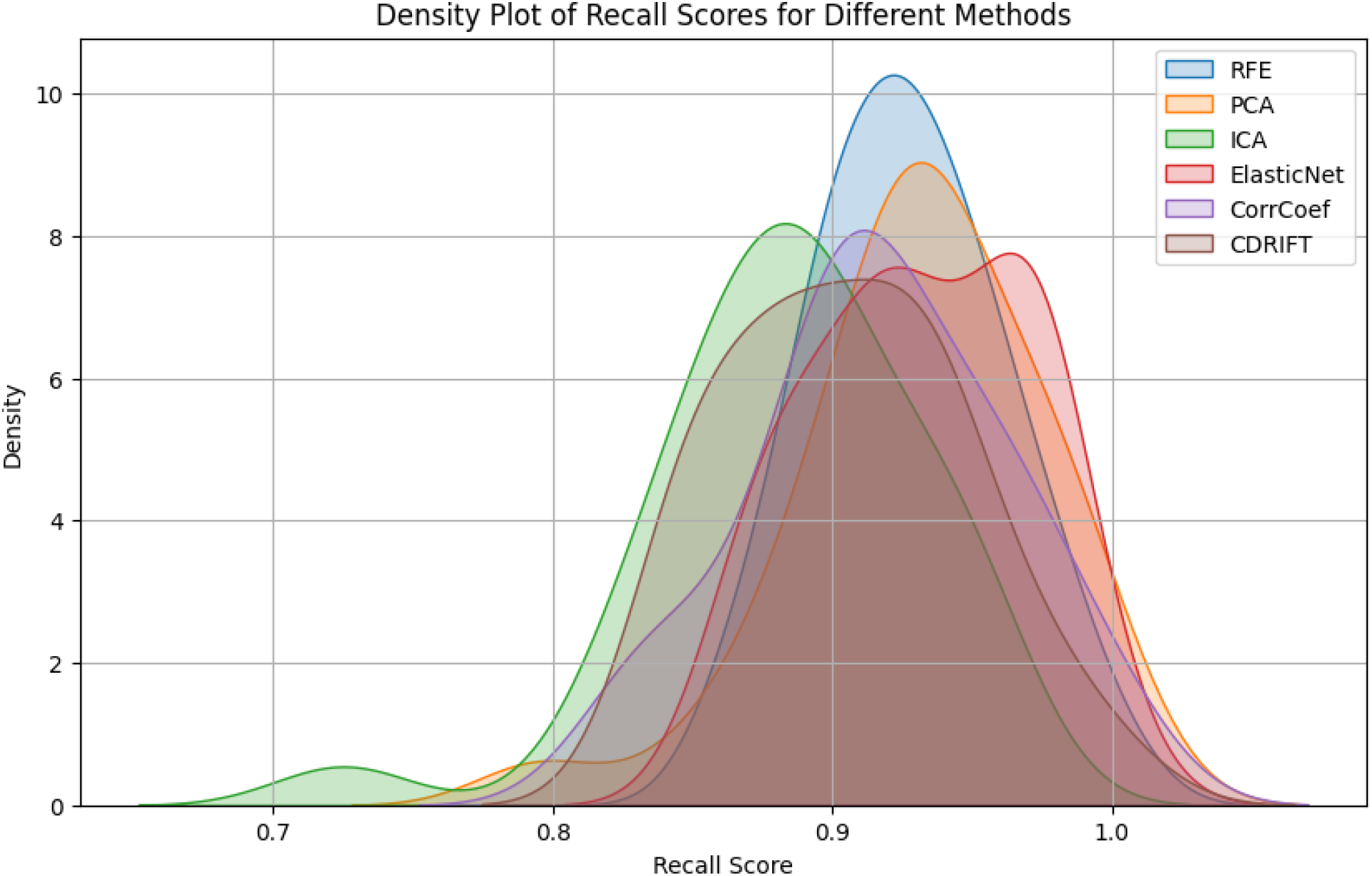
Density plot of recall scores for different methods

Figure 7 visualizes that most methods cluster recall scores between 0.85 and 0.97, indicating generally strong performance. ICA (green curve) shows a noticeable left shift and lower peak, suggesting poorer recall and higher variability than other methods. CausalDRIFT (CDRIFT) has a high, narrow, and symmetrical peak near the top end of the distribution (around 0.93–0.96), again indicating more consistency in performance, this time being strong recall with low variance. RFE, ElasticNet, and PCA also show competitive performance but with slightly wider distributions, implying more variability than CausalDRIFT. CorrCoef performs reasonably but with more spread than the top methods. Therefore, CausalDRIFT offers robust and stable recall performance across repeated runs, while ICA demonstrates significantly lower and more erratic recall, aligning with the ANOVA and Tukey HSD results.

## 6 Conclusion

In this study, we presented CausalDRIFT, a novel causal feature selection algorithm designed to address the limitations of traditional correlation-based methods in healthcare machine learning. By leveraging principles from causal inference, specifically, residualization and Average Treatment Effect (ATE) estimation, CausalDRIFT identifies features with genuine causal influence on clinical outcomes, enhancing model interpretability and robustness. Our experiments across diverse medical datasets (Heart Disease, Diabetes, Breast Cancer, and PCOS) demonstrated that CausalDRIFT achieves competitive predictive performance while prioritizing clinically meaningful features. Notably, it excelled in complex, causally ambiguous settings like PCOS (90% accuracy, F1-score 0.90), matching state-of-the-art methods while offering superior interpretability. Its consistency (low variance in performance) and resistance to confounding further underscore its utility for real-world clinical applications, where stability and transparency are paramount. While CausalDRIFT occasionally traded marginal predictive gains for causal fidelity in correlation-dominated datasets (e.g., Diabetes), its ability to disentangle spurious associations from actionable drivers aligns with the needs of high-stakes healthcare decision-making. The algorithm’s modular design also allows integration with domain knowledge and alternative machine learning models, broadening its applicability. Future work may include scaling CausalDRIFT to larger, multimodal datasets (e.g., EHRs with imaging) and incorporating domain-specific causal graphs could further enhance its precision. Additionally, clinical validation studies are needed to assess its impact on real-world outcomes, such as treatment efficacy or diagnostic accuracy. Furthermore, as CausalDRIFT tends to perform better in HDLSS datasets, future work may explore enhancing CausalDRIFT’s performance on low-dimensional or balanced datasets by integrating hybrid causal-statistical heuristics for robust feature selection. Overall, CausalDRIFT advances the integration of causal reasoning into medical AI, offering a principled, scalable, and interpretable alternative to conventional feature selection. By bridging the gap between statistical association and causal inference, it paves the way for more trustworthy and actionable healthcare machine learning systems.

## Data Availability

The study utilized 4 datasets from Kaggle, access link to the datasets is provided in the GitHub repository of this research, as stated in Experiments section.

https://github.com/SakibHasanSimanto/causalsoap

## References

[1] Bellman, R., & Kalaba, R. (1959). A mathematical theory of adaptive control processes. Proceedings of the National Academy of Sciences, 45 (8), 1288–1290.

[2] Jolliffe, I. T., & Cadima, J. (2016). Principal component analysis: a review and recent developments. Philosophical transactions of the royal society A: Mathematical, Physical and Engineering Sciences, 374 (2065), 20150202.

[3] Oja, E., & Hyvarinen, A. (2000). Independent component analysis: algorithms and applications. Neural networks, 13 (4-5), 411–430.

[4] Chen, X. W., & Jeong, J. C. (2007). Enhanced recursive feature elimination. In Sixth international conference on machine learning and applications (ICMLA 2007) (pp. 429–435). IEEE.

[5] Hall, M. A. (1999). Correlation-based feature selection for machine learning (Doctoral dissertation, The University of Waikato).

[6] Neuberg, L. G. (2003). Causality: models, reasoning, and inference, by Judea Pearl, Cambridge University Press, 2000. Econometric Theory, 19 (4), 675–685.

[7] Schölkopf, B., Locatello, F., Bauer, S., Ke, N. R., Kalchbrenner, N., Goyal, A., & Bengio, Y. (2021). Toward causal representation learning. Proceedings of the IEEE, 109 (5), 612–634.

[8] Baek, K., Draper, B. A., Beveridge, J. R., & She, K. (2002). PCA vs. ICA: A Comparison on the FERET Data Set. In JCIS (pp. 824–827).

[9] Zou, H., & Hastie, T. (2005). Regularization and variable selection via the elastic net. Journal of the Royal Statistical Society Series B: Statistical Methodology, 67 (2), 301–320.

[10] Subbaswamy, A., & Saria, S. (2020). From development to deployment: dataset shift, causality, and shift-stable models in health AI. Biostatistics, 21 (2), 345–352.

[11] Amini, F., & Hu, G. (2021). A two-layer feature selection method using genetic algorithm and elastic net. Expert Systems with Applications, 166, 114072.

[12] Binkyte, R., Sheth, I., Jin, Z., Havaei, M., Schölkopf, B., & Fritz, M. (2025). Causality Is Key to Understand and Balance Multiple Goals in Trustworthy ML and Foundation Models. arXiv preprint 2502.21123.

[13] Stern, A. D., & Price, W. N. (2020). Regulatory oversight, causal inference, and safe and effective health care machine learning. Biostatistics, 21 (2), 363–367.

[14] Glymour, C., Zhang, K., & Spirtes, P. (2019). Review of causal discovery methods based on graphical models. Frontiers in genetics, 10, 524.

[15] Loftus, J. R., Russell, C., Kusner, M. J., & Silva, R. (2018). Causal reasoning for algorithmic fairness. arXiv preprint 1805.05859.

[16] Chernozhukov, V., Chetverikov, D., Demirer, M., Duflo, E., Hansen, C., Newey, W., & Robins, J. (2018). Double/debiased machine learning for treatment and structural parameters. The Econometrics Journal, 21 (1), 1–68.

[17] Hotelling, H. (1933). Analysis of a complex of statistical variables into principal components. Journal of Educational Psychology, 24 (6), 417–441.

[18] Hinton, G. E., & Salakhutdinov, R. R. (2006). Reducing the dimensionality of data with neural networks. Science, 313 (5786), 504–507.

[19] Guyon, I., Weston, J., Barnhill, S., & Vapnik, V. (2002). Gene selection for cancer classification using support vector machines. Machine Learning, 46 (1), 389–422.

[20] Zhang, R., Yang, Q., Wang, X., Wu, H., Zhou, Q., Wang, Y., … & Zhou, F. (2025). DeepSelective: Feature gating and representation matching for interpretable clinical prediction. arXiv preprint 2504.11264.

[21] Mostafa, G., Mahmoud, H., Abd El-Hafeez, T., & E. ElAraby, M. (2024). The power of deep learning in simplifying feature selection for hepatocellular carcinoma: a review. BMC Medical Informatics and Decision Making, 24 (1), 287.

[22] Adnan, K. M., Ghazal, T. M., Saleem, M., Farooq, M. S., Yeun, C. Y., Ahmad, M., & Lee, S. W. (2025). Deep learning driven interpretable and informed decision making model for brain tumour prediction using explainable AI. Scientific Reports, 15 (1), 1–23.

[23] Hai, A. A., Weiner, M. G., Livshits, A., Brown, J. R., Paranjape, A., Hwang, W., … & Rubin, D. J. (2024). Domain generalization for enhanced predictions of hospital readmission on unseen domains among patients with diabetes. Artificial Intelligence in Medicine, 158, 103010.

[24] Nastl, V., & Hardt, M. (2024). Do causal predictors generalize better to new domains? Advances in Neural Information Processing Systems, 37, 31202–31315.

[25] Wu, J., Liu, Z., Zhou, X., Huang, Y., Liu, C., & Lan, C. (2024). Causal Inference-Based Feature Selection Method for Identifying Alzheimer’s Disease Biomarker. In International Conference on Applied Intelligence (pp. 103–114).

[26] Spirtes, P., Glymour, C. N., & Scheines, R. (2000). Causation, prediction, and search. MIT press.

[27] Arjovsky, M., Bottou, L., Gulrajani, I., & Lopez-Paz, D. (2019). Invariant risk minimization. arXiv preprint 1907.02893.

[28] Rothenhäusler, D., Meinshausen, N., Bühlmann, P., & Peters, J. (2021). Anchor regression: Heterogeneous data meet causality. Journal of the Royal Statistical Society Series B: Statistical Methodology, 83 (2), 215–246.

[29] Kwak, S. G., & Kim, J. H. (2017). Central limit theorem: the cornerstone of modern statistics. Korean journal of anesthesiology, 70 (2), 144.

